# Professionalism Pulse: Development and Validation of a Natural Language Processing Pipeline and Dashboard for Safety Culture Surveillance in NYC Health + Hospitals

**DOI:** 10.64898/2026.05.19.26353620

**Authors:** Eunbyul Mangut, Regina Wallace

## Abstract

**Background:** Professionalism and effective communication are foundational determinants of patient safety and quality of care. Unprofessional behaviors frequently serve as active precursors to adverse clinical events. However, proactive organizational surveillance is often hindered because incident feedback exists primarily as unstructured, free-text data. This study aimed to develop and validate a Natural Language Processing (NLP) pipeline and interactive dashboard to proactively monitor the “professionalism climate” within NYC Health + Hospitals, the largest municipal healthcare delivery system in the United States.

**Methods:** A high-fidelity synthetic dataset (N=400) was computationally generated to safely mirror historical incident logs across 11 acute facilities without utilizing Protected Health Information (PHI). A rule-based NLP pipeline was developed in R utilizing the tidytext package. Unstructured narrative feedback was tokenized and classified into three core domains: Respect, Safety, and Communication. To validate the pipeline’s accuracy, a 25% random stratified sample (n=100) was evaluated against independent, blinded manual coding performed by two reviewers, with inter-rater reliability measured via Cohen’s Kappa. Finally, an interactive Tableau dashboard was developed to operationalize and visualize these metrics for ongoing surveillance.

**Results:** The NLP algorithm achieved an overall accuracy of 85.8% (95% CI: 79.0-92.6), with 81.2% sensitivity and 88.9% specificity. The highest domain-specific performance was observed in Communication (88.0% accuracy). Manual validation demonstrated strong inter-rater reliability (k=0.84). Operational analysis via the dashboard revealed that 61.8% of reports occurred during the Tour 2 shift (15:00 to 23:00), aligning with peak operational volume. Furthermore, Respect-related feedback was reported at a disproportionately high frequency during the Tour 3 shift (23:00 to 07:00), accounting for over 50.7% of overnight feedback submissions.

**Conclusion:** Rule-based NLP successfully transforms qualitative healthcare feedback into structured, actionable intelligence with high specificity. Integrating this pipeline into operational dashboards transitions safety culture surveillance from a reactive, manual exercise to a proactive, scalable system, enabling targeted, data-driven interventions by hospital leadership.

## Introduction

The delivery of safe, high-quality patient care is fundamentally dependent upon organizational culture and the professionalism of the clinical workforce. A robust body of literature demonstrates that disruptive behavior, incivility, and poor communication are not merely human resources issues, but active precursors to adverse clinical events. Incivility has been shown to degrade team cognitive function, reduce diagnostic accuracy, and impair the collaborative dynamics necessary for complex medical decision-making (Riskin et al., 2015; Rosen et al., 2018; Katz et al., 2019; Westbrook et al., 2018). Recognizing this critical link, regulatory bodies such as The Joint Commission have mandated that healthcare organizations establish continuous surveillance systems to detect and address “weak signals” of cultural deterioration before they manifest as patient harm (The Joint Commission, 2021).

Despite clear regulatory mandates, healthcare systems often struggle to operationalize safety culture surveillance effectively. Hospitals actively collect vast amounts of staff feedback, peer-reporting data, and incident logs. However, this information constitutes a “data goldmine” that remains largely untapped because it is buried in unstructured, free-text narrative fields. Traditional manual chart abstraction and qualitative review processes are retrospective, highly resource-intensive, slow, and prone to subjective reviewer bias (Cresswell et al., 2020; Kreimeyer et al., 2017; Wu et al., 2022).

Population health informatics provides a solution through Natural Language Processing (NLP). NLP has seen widespread adoption in healthcare for disease prediction and clinical documentation extraction (Wang et al., 2019; Deng & Wang, 2021). However, a significant knowledge gap remains: the vast majority of existing NLP models focus on clinical notes for epidemiological tracking, while very few leverage administrative, HR, or peer-reporting logs for the purpose of prospective culture surveillance.

To address this gap within the context of the largest municipal healthcare system in the United States, this study established three primary objectives. First, to develop an R-based NLP pipeline to automatically categorize unstructured, free-text staff feedback into three core professionalism domains (Respect, Safety, and Communication).

Second, to rigorously validate the pipeline against a human-coded gold standard to ensure the algorithm possesses the high specificity required for operational deployment. Finally, to develop a prototype interactive dashboard (“Professionalism Pulse”) to translate these NLP classifications into rapid, executive-level visual intelligence. By successfully automating the classification of qualitative data, this pipeline aims to shift safety culture surveillance from a reactive reporting system to a proactive, data-driven intervention strategy.

## Methods

### Study Design and Setting

This study utilized a retrospective, cross-sectional analytic design. The project was modeled after the operational environment of NYC Health + Hospitals (NYC H+H), designed to capture the unique scale, linguistic diversity, and departmental characteristics of an expansive, multi-facility public healthcare network.

### Data Sources and Synthetic Generation

Since incident reports and staff feedback logs contain extremely sensitive Protected Health Information (PHI) and confidential Human Resources (HR) data, direct access to live organizational databases was restricted by internal data governance protocols, corporate compliance policies, and the Health Insurance Portability and Accountability Act (HIPAA). To overcome this barrier while maintaining scientific rigor, a high-fidelity synthetic dataset (N=400) was computationally generated. Categorical variables, including the reporting Facility (1-11), Department Involved (e.g., Nursing, Medicine, Surgery), and Operational Shift (Tour 1: 07:00-15:00; Tour 2: 15:00-23:00; Tour 3: 23:00-07:00), were populated using probability distributions that accurately mirror historical volume metrics. To generate the free-text narratives, randomized text templates were populated using standardized clinical vocabulary and established incident typologies. This ensured the dataset maintains high fidelity to the linguistic patterns, lengths, and structures of real-world reporting, without exposing any actual personnel or patient identifiers.

### Variables and Data Pre-processing

The primary independent variables included the operational shift, department, and facility. The dependent variable was the assigned Professionalism Domain (Respect, Safety, Communication). Data cleaning and pre-processing were conducted using R software (version 4.3.1), heavily relying on the tidytext (v0.4.1) and dplyr (v1.1.2) packages. The raw narrative text underwent normalization, which included lowercasing all characters and removing punctuation and special symbols, followed by stop-word filtration to remove common, non-informative English words (e.g., “the,” “and,” “is”). Subsequently, the text underwent tokenization. Tokenization is a foundational computational process in text mining that dissects continuous strings of text (sentences) into smaller, discrete units, specifically individual words (unigrams) and two-word phrases (bigrams). By breaking the text down into these tokens, the algorithm could perform quantitative frequency analysis and match the narrative against a predefined lexicon (see Figure 1).

**Figure 1.**
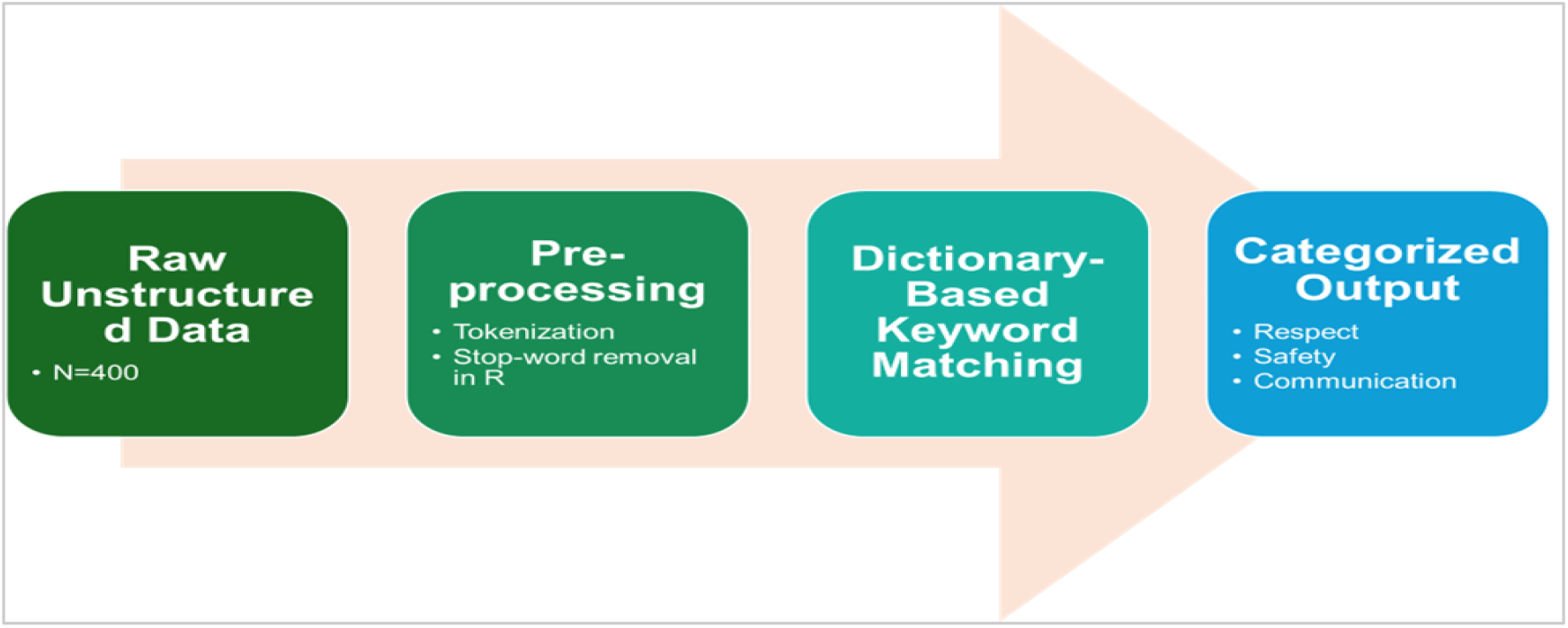
Natural Language Processing (NLP) Pipeline Workflow. Graphical summary of the automated text-mining pipeline developed in R. Raw unstructured feedback text undergoes pre-processing (lowercasing, punctuation removal, stop-word exclusion) and tokenization. A dictionary-based lexicon matches unigram and bigrams against a predefined taxonomy, classifying records into three mutually exclusive professionalism domains (Safety, Respect, Communication) for subsequent visualization in the Tableau dashboard.

### NLP Pipeline and Statistical Analysis

A rule-based, dictionary-matching NLP pipeline was constructed. A customized lexicon was developed, assigning specific clinical and behavioral keywords to the Respect, Safety, and Communication domains. The algorithm reviewed the tokenized narratives and assigned a primary domain classification based on keyword density. Descriptive statistics (counts, percentages, means) were calculated to summarize the cohort. Algorithm performance was evaluated using standard diagnostic metrics: Accuracy, Sensitivity, Specificity, and 95% Confidence Intervals (CIs). Finally, an interactive dashboard was developed using Tableau Desktop software (version 2023.2) to operationalize and visualize these metrics for ongoing surveillance.

### Validation Strategy and Ethical Considerations

To establish a “gold standard” for algorithm validation, a 25% random sample (n=100) was extracted for manual review. Independent, blinded manual coding was performed by two distinct reviewers: the primary investigator (E.M.) and a subject matter expert overseeing continuous professional education (R.W.). To assess inter-rater reliability (IRR) between the two human coders, a 20% subset of the validation records (n=20) was double-coded and evaluated using Cohen’s Kappa (k). The study utilized entirely synthetic data; therefore, it did not qualify as human subjects research requiring Institutional Review Board (IRB) approval.

## Results

### Descriptive Statistics

The synthetic cohort (N=400) accurately reflected a diverse, multi-facility healthcare system. The highest volume of feedback reports originated from the Nursing department (22.3%), followed by Medicine (19.3%). Registered Nurses (RNs) represented the dominant reporting role, accounting for 49.0% of all submitted staff feedback (Table 1).

**Table 1.** Baseline Characteristics of the Unstructured Staff Feedback Dataset (N=400). Demographic and operational characteristics of 400 synthetic staff feedback records simulating the NYC Health + Hospitals reporting environment from January 2025 to December 2025. Categorical variables are presented as frequency and percentage. The dataset assumes a roughly uniform distribution across facilities, shift, and roles to simulate baseline institutional reporting prior to NLP categorization. Specific NYC H+H facility names have been blinded (Facility 1-11) for presentation purposes. Abbreviations: NYC H+H, New York City Health + Hospitals; PA, Physician Assistant; PCA, Patient Care Assistant; RN, Registered Nurse.

| Characteristic | Total Cohort (N=400) | Percentage (%) |
| --- | --- | --- |
| Facility Setting |  |  |
| Facility 1 | 68 | 17.0% |
| Facility 2 | 51 | 12.8% |
| Facility 3 | 39 | 9.8% |
| Facility 4 | 21 | 5.3% |
| Facility 5 | 35 | 8.8% |
| Facility 6 | 31 | 7.8% |
| Facility 7 | 48 | 12.0% |
| Facility 8 | 33 | 8.3% |
| Facility 9 | 9 | 2.3% |
| Facility 10 | 53 | 13.3% |
| Facility 11 | 12 | 3.0% |
| **Department Involved** |  |  |
| Administration | 37 | 9.3% |
| Ambulatory Care | 12 | 3.0% |
| Critical Care | 10 | 2.5% |
| Dietary | 7 | 1.8% |
| Emergency Department | 60 | 15.0% |
| Environmental Services | 11 | 2.8% |
| Labor and Delivery | 5 | 1.3% |
| Medicine | 77 | 19.3% |
| Nursing | 89 | 22.3% |
| OB/ GYN | 5 | 1.3% |
| Pediatrics | 21 | 5.3% |
| Pharmacy | 4 | 1.0% |
| Psychology/ Psychiatry | 15 | 3.8% |
| Radiology | 8 | 2.0% |
| Rehabilitation Medicine | 5 | 1.3% |
| Respiratory Therapy | 4 | 1.0% |
| Social Work | 12 | 3.0% |
| Surgery | 18 | 4.5% |
| **Operational Shift (Author Tour)** |  |  |
| Tour 1 (07:00-15:00) | 86 | 21.5% |
| Tour 2 (15:00-23:00) | 247 | 61.8% |
| Tour 3 (23:00-07:00) | 67 | 16.8% |
| **Reporting Role (Author Title)** |  |  |
| Administration | 14 | 3.5% |
| Attending Physician | 8 | 2.0% |
| Clerical Associate | 16 | 4.0% |
| Head Nurse | 8 | 2.0% |
| Nursing Administrator | 13 | 3.3% |
| Other | 13 | 3.3% |
| PA | 4 | 1.0% |
| PCA | 12 | 3.0% |
| Pharmacist | 5 | 1.3% |
| Resident | 87 | 21.8% |
| RN | 196 | 49.0% |
| Social Worker | 24 | 6.0% |

### Primary Findings: Algorithm Validation and Scope

When tested against the human-coded gold standard, the NLP algorithm demonstrated strong classification capabilities. The pipeline achieved an overall average accuracy of 85.8% (95% CI: 79.0-92.6) (see Table 2). Performance was highest within the Communication domain, which reached an accuracy of 88.0%. The pipeline prioritized high specificity, achieving an average of 88.9%. This indicates that the algorithm successfully ignored irrelevant text and accurately identified true negatives, which is crucial for minimizing “false alarms” in operational surveillance. Conversely, the algorithm’s sensitivity was recorded at 81.2%. This metric defines the scope and coverage of the tool, indicating a false-negative rate of approximately 18.8%. In practical terms, nearly one in five reports containing professionalism issues were missed by the algorithm. Analysis of these false negatives revealed that they typically involved complex clinical jargon, novel slang, or implicit, highly nuanced descriptions of incivility that did not trigger the matching algorithms of the rule-based dictionary. Inter-rater reliability between the two human coders for the validation set was strong, yielding a Cohen’s k of 0.84, confirming that the ground-truth standard was rigorous and consistent.

**Table 2.** Preliminary Validation Metrics of the Rule-Based NLP Algorithm (n=100 subset). Performance metrics of the R-based classification algorithm against a manually coded “gold standard” subset of 100 randomly selected records. True positives, false positives, true negatives, and false negatives were calculated for each domain independently. Accuracy represents the overall proportion of correct classifications. Sensitivity indicates the algorithm’s ability to correctly identify a specific professionalism issue, while specificity indicates the ability to correctly identify true negative records.

| Professionalism Domain | Accuracy (%)<br>(95% CI) | Sensitivity (%)<br>(95% CI) | Specificity (%)<br>(95% CI) |
| --- | --- | --- | --- |
| Safety | 86.0<br>(79.2-92.8) | 78.5<br>(64.5-92.5) | 91.2<br>(84.4-98.0) |
| Respect | 83.5<br>(76.2-90.8) | 81.0<br>(67.6-94.4) | 85.0<br>(76.5-93.5) |
| Communication | 88.0<br>(81.6-94.4) | 84.2<br>(71.8-96.6) | 90.5<br>(83.5-97.5) |
| Overall Model Average | 85.8<br>(79.0-92.6) | 81.2<br>(73.6-88.8) | 88.9<br>(84.6-93.2) |

### Secondary Findings: Dashboard and Operational Insights

The classified NLP outputs were successfully ingested into a Tableau dashboard (“Professionalism Pulse”) to visualize trends. The dashboard revealed significant temporal variations in reporting behaviors. Notably, 61.8% of all feedback was generated during Tour 2 (15:00 to 23:00), aligning closely with the peak operational hours of patient and discharge cycles. Within this high-volume period, Respect-related feedback submissions were the primary driver of reports, indicating the interpersonal pressure of peak-flow environments. Furthermore, a drill-down analysis of the domains presented a disproportionate spike in the concentration of these issues during Tour 3 (23:00 to 07:00), where Respect accounted for over 50.7% of overnight reports (Figure 2). This highlights a specific vulnerability during the overnight shift, suggesting that while the afternoon requires support for high volume, the overnight shift requires targeted interventions to address interpersonal professional conduct. Further demographic drilldowns demonstrated these high-volume reporting trends remained consistent across specific clinical departments (Figure 3) and frontline professional roles (Figure 4).

**Figure 2.**
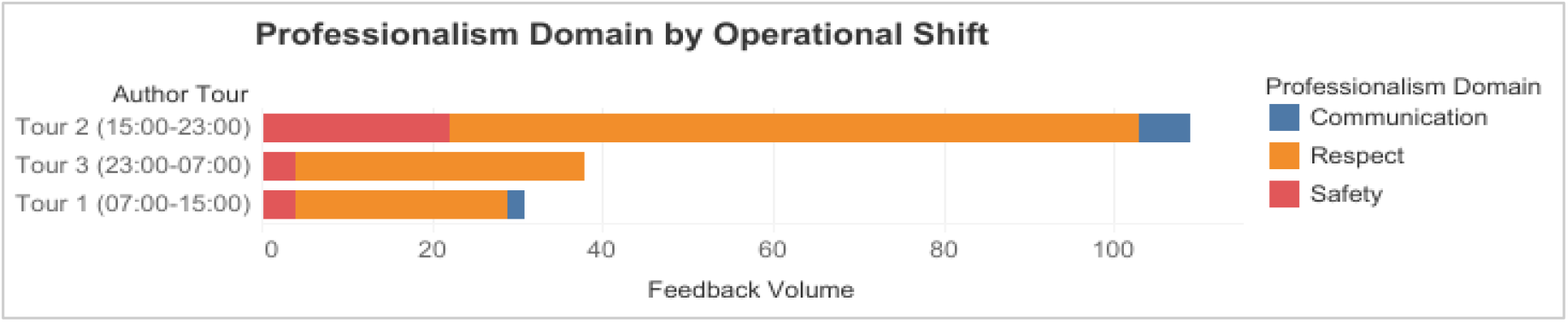
Dashboard Chart: Professionalism Domain by Operational Shift. Visualization of NLP-categorized incident reports across three standard operational shifts. Categorical comparisons are presented as absolute frequencies. Trend analysis indicates Respect-related feedback peaks disproportionately during Tour 3, potentially correlating with reduced nighttime administrative oversight and staffing ratios. This serves as the foundational view for the “Professionalism Pulse” dashboard.

**Figure 3.**
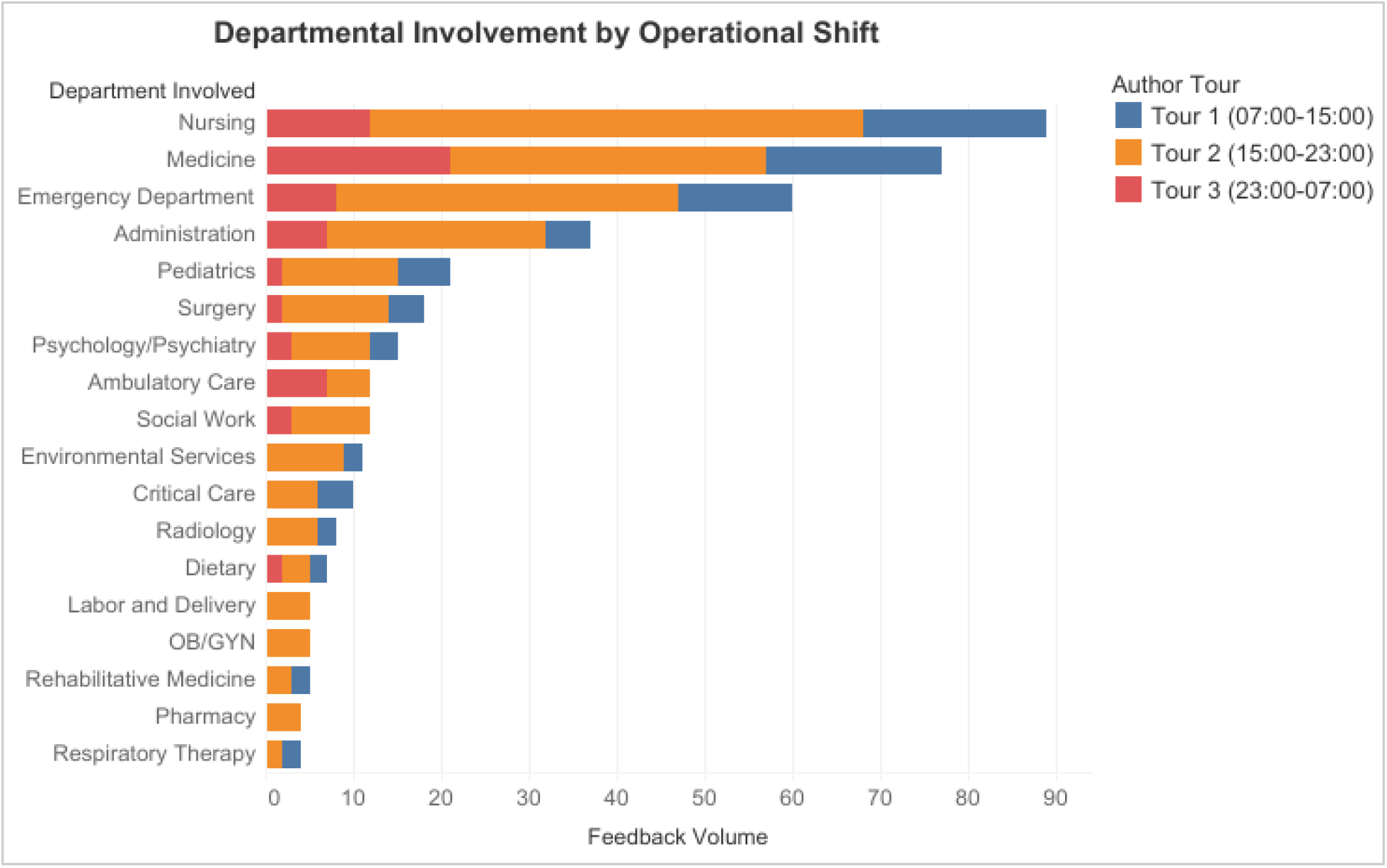
Dashboard Chart: Department Involvement by Operational Shift. Visualization of NLP-categorized incident reports stratified by clinical and administrative departments across three operational shifts. Categorical comparisons are presented as absolute frequencies. Trend analysis indicates Nursing, Medicine, and Emergency departments generate the highest feedback volumes, peaking during Tour 2, likely due to high afternoon patient turnover. This serves as a secondary demographic drill-down for the dashboard.

**Figure 4.**
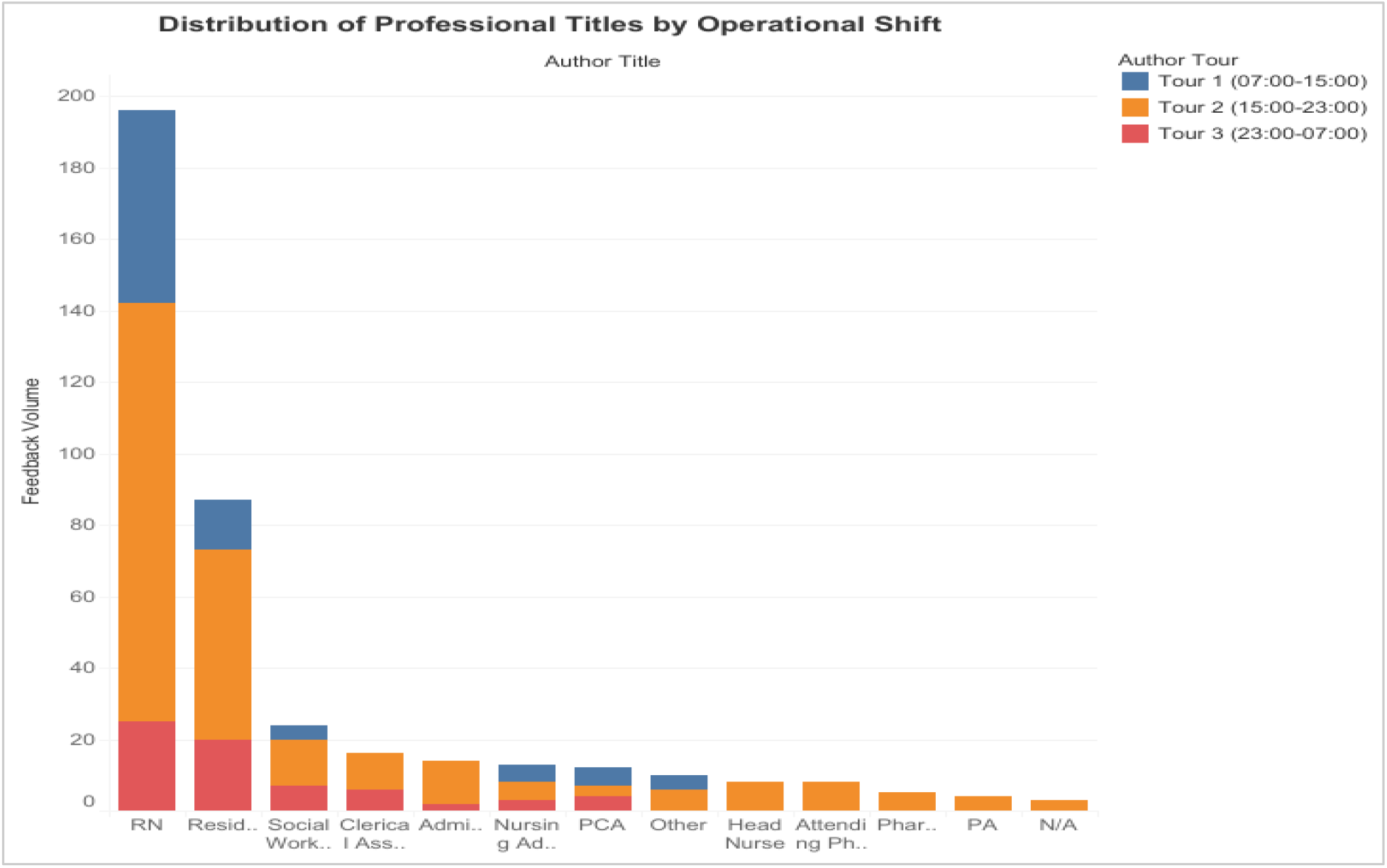
Dashboard Chart: Professional (Author) Titles by Operational Shift. Visualization of NLP-categorized incident reports stratified by professional author title across three operational shifts. Categorical comparisons are presented as absolute frequencies. Trend analysis indicates Registered Nurses (RNs) and Residents generate the highest feedback volumes, peaking during Tour 2, reflecting their sustained frontline clinical exposure. The “N/A” category represents feedback submitted anonymously where the reporting professional’s title was omitted. This serves as a tertiary demographic drill-down for the dashboard.

## Discussion

This study successfully demonstrated that unstructured, qualitative healthcare staff feedback can be reliably transformed into structured, quantitative intelligence using Natural Language Processing. The high overall accuracy (85.8%) and excellent specificity (88.9%) prove that the algorithm is highly effective at filtering out irrelevant noise, a vital necessity for operational deployment. Furthermore, integrating these structured outputs into a visualization dashboard allowed for the immediate identification of temporal and operational anomalies, such as the concentrated Respect-related risks observed during the Tour 3 night shift.

These findings align with emerging literature indicating that informatics tools are the critical bridge between frontline data collection and executive action (Fong et al., 2017). Previous studies by Leape et al. (2012) emphasized that addressing disrespect requires a reliable system of accountability; however, measuring that accountability has historically been subjective. By automating the categorization of incident logs, this pipeline addresses the operational bottleneck cited by Deng & Wang (2021), proving that administrative logs hold as much predictive value for safety culture as clinical documentation does for disease management.

A primary strength of this study is its highly rigorous validation methodology. By utilizing two distinct human coders and achieving strong inter-rater reliability (k=0.84), this study ensured the algorithm was judged against an unbiased ground truth. Furthermore, the innovative use of synthetic data allowed scalable, computationally reproducible algorithm development without risking patient privacy or violating HIPAA regulations. However, several limitations must be reviewed. As quantified by the 81.2% sensitivity rate, the algorithm’s scope is inherently restricted by its rule-based architecture. Dictionary-matching NLP struggles to comprehend contextual sarcasm, severe typographical errors, or newly adopted clinical jargon, leading to a notable volume of false negatives. Additionally, since the study relied on synthetic data, immediate, live operational interventions could not be deployed based on these specific numerical findings.

The public health implications of this tool are significant. By replacing weeks of manual, labor-intensive feedback reviews with real-time dashboard metrics, hospital leadership can deploy targeted, unit-level interventions precisely when and where they are needed. Future research should focus on algorithm maturity (Khairat et al., 2018; Dowding et al., 2015). Upgrading the pipeline from a rule-based dictionary framework to an advanced machine learning model, such as a Large Language Model (LLM) or a specialized BERT architecture, would greatly improve the algorithm’s sensitivity and its ability to understand nuanced clinical context. Additionally, future efforts must focus on securely integrating this architecture directly into live IT infrastructure to close the feedback loop, linking the dashboard’s insights directly to Continuing Professional Education (CPE) initiatives to track subsequent reductions in adverse incidents.

## Conclusion

The “Professionalism Pulse” pipeline demonstrates that it is computationally feasible to proactively monitor a healthcare organization’s safety culture. By leveraging NLP and data visualization, health systems can unlock the hidden value in their qualitative data, moving beyond retrospective damage control. By actively fostering a culture of respect and operational excellence, healthcare organizations can directly translate administrative intelligence into sustained improvements in patient safety and overall quality of care.

## Data Availability

The dataset analyzed during the current study is entirely synthetic and computationally modeled. However, to comply with organizational data governance and internal risk management protocols, the full dataset and its underlying generation parameters are restricted and not available for public distribution.

## Appendix A Interactive Dashboard Prototype

**Figure A1.**
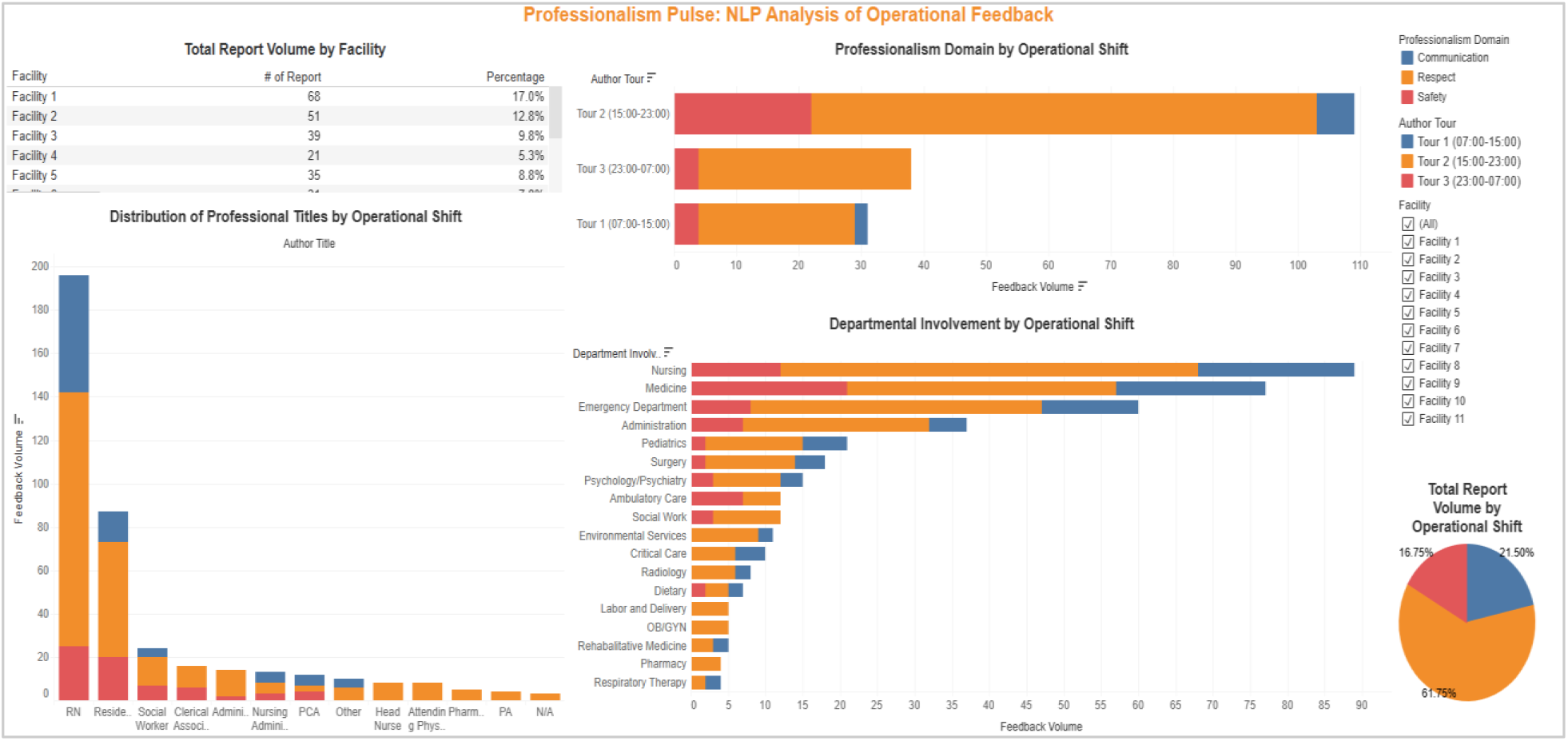
Professionalism Pulse: Executive Surveillance Dashboard. Full-scale prototype of the Tableau “Professionalism Pulse” interactive dashboard. This executive view aggregates NLP-classified staff feedback (N=400) to enable proactive safety culture surveillance. The interface features dynamic filtering capabilities by reporting facility, temporal tracking, and high-level professionalism domain distribution metrics to support targeted, data-driven interventions by hospital leadership.

## Appendix B Natural Language Processing (NLP) Rule-Based Lexicon Taxonomy

**Table B1.** Sample Taxonomy of NLP Lexicon by Professionalism Domain. Sample taxonomy of clinical and behavioral unigrams and bigrams utilized by the rule-based NLP algorithm. These tokens were used to train the customized dictionary, allowing the pipeline to automatically classify unstructured, free-text narrative feedback into one of three mutually exclusive primary professionalism domains.

| Safety | Respect | Communication |
| --- | --- | --- |
| patient fall | yelled at | handoff error / issue |
| medication error | rude tone | unclear instructions |
| slip / trip | condescending | delayed response |
| hazard | ignored | page ignored |
| needle stick | unprofessional | not informed |
| wrong dose / route | hostile | missing documentation |
| broken equipment | insulted | language barrier |
| spill | dismissive | misunderstood |
| unsafe practice | bothered | failed to notify |
| injury | inappropriate language | missing information |
| infection control | abusive | unresponsiveness |
| protocol failure / breach | raised voice | poor sign-out |

## Appendix C Synthetic Data Generation Parameters

**Table C1.** Baseline Probability Distribution for Synthetic Dataset Generation (N=400). Baseline probability distribution parameters utilized to computationally generate the high-fidelity synthetic dataset prior to NLP analysis. Weights were modeled after historical reporting volumes and workforce demographics to ensure structural validity and realistic organizational representation without utilizing actual Protected Health Information (PHI) or confidential Human Resources (HR) data.

| Variable Category | Categorical Level | Assigned Probability Weight |
| --- | --- | --- |
| Operational Shift | Tour 2 (15:00-23:00) | 0.618 |
|  | Tour 1 (07:00-15:00) | 0.215 |
|  | Tour 3 (23:00-07:00) | 0.168 |
| Author Title | Registered Nurse (RN) | 0.490 |
|  | Resident Physician | 0.218 |
|  | Social Worker | 0.060 |
|  | Clerical / Administration | 0.075 |
|  | Nursing Leadership | 0.053 |
|  | Patient Care Assistant (PCA) | 0.030 |
|  | Attending Physician | 0.020 |
|  | Other Staff | 0.055 |
| Department | Nursing | 0.223 |
|  | Medicine | 0.193 |
|  | Emergency Department | 0.150 |
|  | Administration | 0.093 |
|  | Pediatrics | 0.053 |
|  | Surgery | 0.045 |
|  | Psychiatric / Psychology | 0.038 |
|  | Other Clinical / Ancillary | 0.208 |

